# History of episodic heavy alcohol use predicts antidepressant effectiveness of ketamine

**DOI:** 10.1101/2025.08.09.25333370

**Authors:** Aaishah R. Raquib, Vy Nguyen, Joshua C. Brown, Amanda E. Sedgewick, Stephen Seiner, Joseph J. Taylor, Shan H. Siddiqi, Robert C. Meisner

## Abstract

**Rationale:** Antidepressant response to ketamine is greatest in patients with a family history of alcohol use disorder (AUD), but it remains unclear if this trend holds true for a personal history of alcohol use.

**Objective:** We hypothesized that a personal history of episodic heavy drinking would positively correlate with antidepressant response to ketamine.

**Methods:** In a retrospective cohort study, patients with Major Depressive Disorder (MDD) received intravenous ketamine. To assess specificity, patients receiving electroconvulsive therapy (ECT) at the same center were also studied. Depression was quantified using the Quick Inventory of Depressive Symptomatology (Self-Report) (QIDS-SR), and alcohol use was quantified using the Alcohol Use Disorder Identification Test question 3 (AUDIT-3).

**Results:** Of 225 patients evaluated for ketamine, 178 completed the AUDIT-3 and completed the initial course of three treatments. Of 2577 that received ECT, 1851 received at least 10 treatments. Across both groups, patients were between age 16 and 90. With ketamine, history of past-year episodic heavy drinking was associated with greater improvement in QIDS-SR with a significant Group x Time interaction after the first three treatments (F(12,502)=2.25, p=0.0089), four treatments (F(16,632)=2.56, p=0.0007), and five treatments (F(20,757)=2.22, p=0.0016). This effect was not seen with ECT after 5 treatments (F(4,2283)=0.91, p=0.46), 10 treatments (F(8,3939)=1.63, p=0.11), or 20 treatments (F(16,7154)=1.50, p=0.09). There was an interaction after 15 treatments (F(12,5113)=1.93, p=0.027), although this did not survive multiple comparisons correction.

**Conclusions:** In patients with trMDD, history of episodic heavy drinking predicted antidepressant effectiveness of ketamine, but not ECT. These results may assist in patient selection for ECT vs ketamine.

**Funding:** The study was unfunded.

**Competing Interests:** The authors have no relevant conflicts of interest to disclose

## Introduction

Sub-anesthetic doses of ketamine are an effective treatment for some individuals with treatment-resistant major depressive disorder (trMDD), and a family history of alcohol use disorder (AUD) appears to be a positive predictor of response (Petrakis et al. 2004; Niciu et al. 2014). This response pattern may be driven by overlapping mechanisms, as ketamine and alcohol are both N-methyl-D-aspartate (NMDA) receptor antagonists (Petrakis et al. 2004; Phelps et al. 2009) and also both have secondary opioidergic mechanisms (Williams et al. 2018; Avery 2022). Family history of AUD greatly increases a patient’s own risk for developing AUD, which in turn, increases the risk of MDD (Grant 1998; Boden and Fergusson 2011; MacKillop et al. 2022). Patients with AUD are often excluded from ketamine studies, however, so it remains unclear how excessive use of alcohol, or a history thereof, may affect potential effectiveness of ketamine treatment. When assessing an individual’s candidacy for ketamine, or other treatments for trMDD, more guidance is needed for clinicians to evaluate clinical and biological factors that favor one intervention over the other for a given patient.

To further investigate the relationship between alcohol use and the effectiveness of sub-anesthetic treatment with ketamine, we analyzed the relationship between history of episodic heavy drinking and improvement in depressive symptoms over time. To assess specificity to ketamine, we performed the same analysis in patients receiving electroconvulsive therapy (ECT). This specificity analysis is particularly relevant in context of two recent large clinical trials comparing ECT to ketamine directly. Although a perfect trial is not possible due to lack of blinding, one trial showed that ECT is superior to ketamine (Ekstrand et al. 2022), while another found that ketamine is non-inferior for trMDD without psychosis (Anand et al. 2023). Notwithstanding the debate about relative efficacy, this raises questions about which patients are most appropriate for which treatment (Anand et al. 2023). In this study, we sought to determine if episodic heavy drinking in the past year may serve as a marker for personalized medicine in interventional psychiatry for patients with trMDD.

## Methods

Data from individuals who received intravenous ketamine at McLean Hospital before December 2021 were collected in a REDCap database as part of routine clinical care. This database was retrospectively analyzed in a deidentified manner, with the approval of the Mass General Brigham Institutional Review Board. To assess whether the effect is specific to ketamine, we also included patients receiving ECT at McLean Hospital before December 2021.

IV racemic ketamine was first offered in April 2018, while ECT records date back to April 2011. Data for each patient, including treatment dates, Quick Inventory of Depressive Symptomatology (Self-Report) (QIDS-SR) scores at each treatment interval, and Alcohol Use Disorder Identification Test question 3 (AUDIT-3) were extracted from a REDCap database. QIDS-SR was collected immediately before initiation of the ketamine treatment series, and immediately before each treatment to capture interval changes at each treatment encounter. QIDS-SR was collected every 5 treatments with ECT. The QIDS-SR scores were treated as a continuous variable rather than choosing a specific cutoff for response. The AUDIT-3 score, collected once at baseline, was used as our metric for “episodic heavy alcohol use” and only specified drinking habits from the twelve months before self-reporting. Both treatments were administered naturalistically, with doses and treatment parameters titrated based on tolerability and efficacy according to clinical judgment.

Ketamine was administered to treatment-seeking patients who were referred by an outpatient psychiatrist for trMDD. Patients with active substance use disorders were excluded from treatment. Standard clinical practice in the ketamine service included a 3-4 infusion “Challenge” phase to determine responder status. The dose was started at 0.5 mg/kg and was titrated at the clinician’s discretion, up to a limit of 1.1 mg/kg. After the Challenge phase, patients demonstrating substantive responses on objective scales and subjective psychiatric evaluation continued to complete 8 total infusions. Substantive response was defined based on multifactorial clinical assessment, not based on specific scores on the QIDS-SR. Patients who demonstrated no substantive response after the Challenge Phase discontinued treatment. For the present analysis, treatment outcomes were considered after three, four, and five infusions. This range was chosen because at least three infusions were conducted in the challenge phase for most participants, and most patients experience benefit with ketamine within 4-5 infusions. Further time points were not included due to risk of selection bias, as patients who remit after 3-5 infusions may be less likely to receive a sixth infusion, while patients who experience no benefit after 3-5 infusions may also be less likely to receive a sixth infusion.

ECT was delivered with a Mecta machine (either Spectrum 5000 or Sigma -Tualatin, OR). Most patients were started using ultrabrief pulse right unilateral ECT three times a week, using a dose titration at the first treatment. Subsequent treatments were started at six times the seizure threshold and adjusted clinically according to treatment response and seizure quality. In cases of inadequate response, some patients were switched to brief pulse unilateral ECT or bifrontal or bilateral placement depending on urgency, tolerability, and severity of illness. Inadequate response was defined based on multifactorial clinical assessment, not based on specific scores on the QIDS-SR. Patients with current or past history of alcohol use disorder were allowed to participate in treatment if they agreed to remain sober for the treatment period. Significant relapses during treatment usually resulted in discontinuation of treatment to focus on sobriety.

The QIDS-SR was used as a quantitative marker to compare mood between treatment sessions. It was administered every 5 treatments for monitoring of outcomes as part of routine clinical practice. For the present analysis, outcomes were analyzed at the first time point (5 treatments) to parallel the ketamine analysis, and were also measured at subsequent time points (10, 15, and 20 treatments) to account for delayed response, which often occurs with ECT.

The AUDIT-3 question, “How often in the last year have you had 4 or more drinks on one occasion?” for women and “6 or more drinks” for men, was collected once at the baseline visit. A score of 0 corresponds to “never,” 1 is “less than monthly,” 2 is “monthly,” 3 is “weekly”, and 4 is “daily or almost daily.” This question alone has been used as a quantitative screening tool that is validated as a predictor for AUD and heavy drinking in both women (Bradley et al. 2003) and men, and is consistent with previous findings that 5+ drinks on any occasion within the past year predict alcohol use disorder (Dawson 1994; Buchsbaum et al. 1995; Bush et al. 1998; Bradley et al. 2003). For the primary analysis, patients were grouped according to AUDIT-3 score, with each score treated as a different group. For a secondary analysis, AUDIT-3 score was binarized as either zero or greater than zero. Repeated-measures two-way ANOVA was used to assess a group x time interaction on QIDS-SR score. We hypothesized that a history of heavier drinking in the past year would be associated with greater antidepressant response to ketamine, but not to ECT.

## Results

Altogether, 225 patients in the ketamine group and 2,577 patients in the ECT group had at least one follow-up assessment and were thus included. Patients receiving ECT had significantly greater baseline depression severity and significantly higher AUDIT-3 score indicating more frequent heavy drinking (Table 1). Twenty-four percent of patients in the ketamine group (n=53) and 29% of patients in the ECT group (n=757) reported a score of greater than zero, which corresponds to at least one episode of heavy drinking in the past year.

**Table 1:** Baseline clinical characteristics.

|  | **ECT (n = 2577)** | **Ketamine (n= 225)** | **p** |
| --- | --- | --- | --- |
| **Age ± SD** | 45.2 ± 16.8 | 44.5 ± 17 | 0.60 |
| **Gender** | 55% female<br>43% male<br>1% other<br>1% unknown | 54% female<br>43% male<br>3% other<br>1% unknown | 0.04 |
| **Race/ethnicity**<br>(multiple selections allowed) | 89% White<br>1% Native American<br>3% Asian/Pacific Islander<br>2% Black<br>3% Latino<br>2% Other | 92% White<br>1% Native American<br>6% Asian/Pacific Islander<br>0.5% Black<br>2% Latino<br>2% Other | n/a |
| **Time point** | Baseline (time 0): n=2,588<br>Treatment 5: n= 2,350<br>Treatment 10: n= 1,851<br>Treatment 15: n= 1,357 | Baseline (time 0): n= 225<br>Treatment 1: n= 195<br>Treatment 2: n= 197<br>Treatment 3: n= 178<br>Treatment 4: n= 150<br>Treatment 5: n= 152 | n/a |
| **Average AUDIT-3 score** | 0.52 ± 0.94 | 0.35 + 0.70 | 0.002 |
| **AUDIT-3 subgroups** | Score 0: 74%<br>Score 1: 20%<br>Score 2: 4%<br>Score 3: 1%<br>Score 4: 1% | Score 0: 69%<br>Score 1: 19%<br>Score 2: 6%<br>Score 3: 4%<br>Score 4: 2% | 0.142 |
| **Average baseline QIDS-SR score** | 22.31 ± 5.96 | 19.82 ± 6.47 | 6.4 x 10<sup>-8</sup> |

**Table 2:**
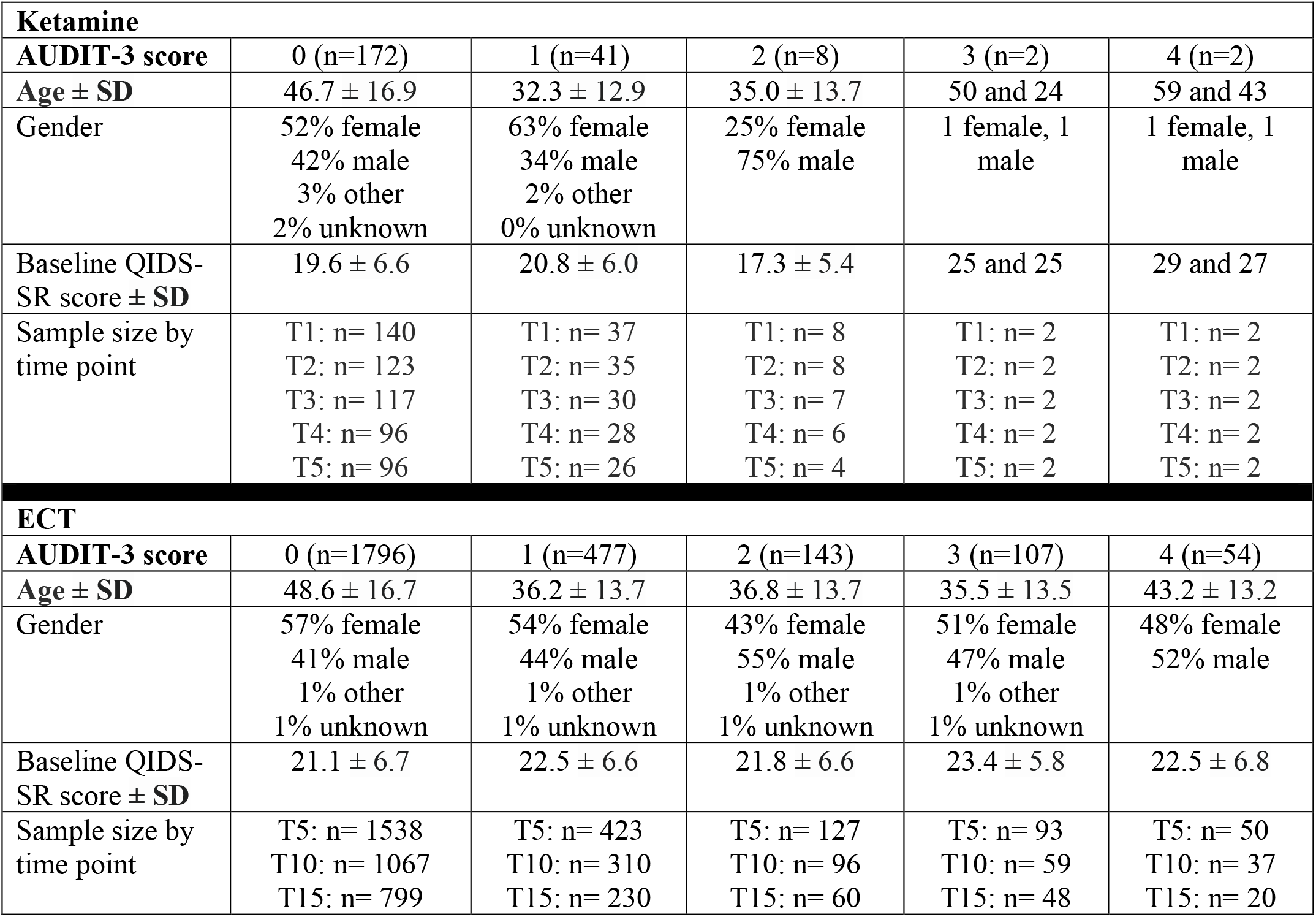
Demographics by AUDIT-3 subgroup.

| **Ketamine** |  |  |  |  |  |
| --- | --- | --- | --- | --- | --- |
| **AUDIT-3 score** | 0 (n=172) | 1 (n=41) | 2 (n=8) | 3 (n=2) | 4 (n=2) |
| **Age ± SD** | 46.7 ± 16.9 | 32.3 ± 12.9 | 35.0 ± 13.7 | 50 and 24 | 59 and 43 |
| **Gender** | 52% female<br>42% male<br>3% other<br>2% unknown | 63% female<br>34% male<br>2% other<br>0% unknown | 25% female<br>75% male | 1 female, 1 male | 1 female, 1 male |
| **Baseline QIDS-SR score ± SD** | 19.6 ± 6.6 | 20.8 ± 6.0 | 17.3 ± 5.4 | 25 and 25 | 29 and 27 |
| **Sample size by time point** | T1: n= 140<br>T2: n= 123<br>T3: n= 117<br>T4: n= 96<br>T5: n= 96 | T1: n= 37<br>T2: n= 35<br>T3: n= 30<br>T4: n= 28<br>T5: n= 26 | T1: n= 8<br>T2: n= 8<br>T3: n= 7<br>T4: n= 6<br>T5: n= 4 | T1: n= 2<br>T2: n= 2<br>T3: n= 2<br>T4: n= 2<br>T5: n= 2 | T1: n= 2<br>T2: n= 2<br>T3: n= 2<br>T4: n= 2<br>T5: n= 2 |
| **ECT** |  |  |  |  |  |
| **AUDIT-3 score** | 0 (n=1796) | 1 (n=477) | 2 (n=143) | 3 (n=107) | 4 (n=54) |
| **Age ± SD** | 48.6 ± 16.7 | 36.2 ± 13.7 | 36.8 ± 13.7 | 35.5 ± 13.5 | 43.2 ± 13.2 |
| **Gender** | 57% female<br>41% male<br>1% other<br>1% unknown | 54% female<br>44% male<br>1% other<br>1% unknown | 43% female<br>55% male<br>1% other<br>1% unknown | 51% female<br>47% male<br>1% other<br>1% unknown | 48% female<br>52% male |
| **Baseline QIDS-SR score ± SD** | 21.1 ± 6.7 | 22.5 ± 6.6 | 21.8 ± 6.6 | 23.4 ± 5.8 | 22.5 ± 6.8 |
| **Sample size by time point** | T5: n= 1538<br>T10: n= 1067<br>T15: n= 799 | T5: n= 423<br>T10: n= 310<br>T15: n= 230 | T5: n= 127<br>T10: n= 96<br>T15: n= 60 | T5: n= 93<br>T10: n= 59<br>T15: n= 48 | T5: n= 50<br>T10: n= 37<br>T15: n= 20 |

Antidepressant efficacy of ketamine was positively associated with heavy episodic alcohol use in the past year, as evidenced by a significant group x time interaction on QIDS-SR scores after the first three treatments (F(12,502)=2.25, p=0.0089), four treatments (F(16,632)=2.56, p=0.0007), and five treatments (F(20,757)=2.22, p=0.0016) (Fig. 1a). Similar results were observed when binarizing alcohol use based on AUDIT-3 score of zero (no episodes of excessive drinking in the past year) or greater than zero (1+ episode(s) of excessive drinking) – a significant group x time interaction was observed after three treatments (F(3,512)=3.46, p=0.016), four treatments (F(4,645)=5.18, p=0.00041), or five treatments (F(5,774)=4.69, p=0.0003) (Fig. 1b).

**Figure 1:**
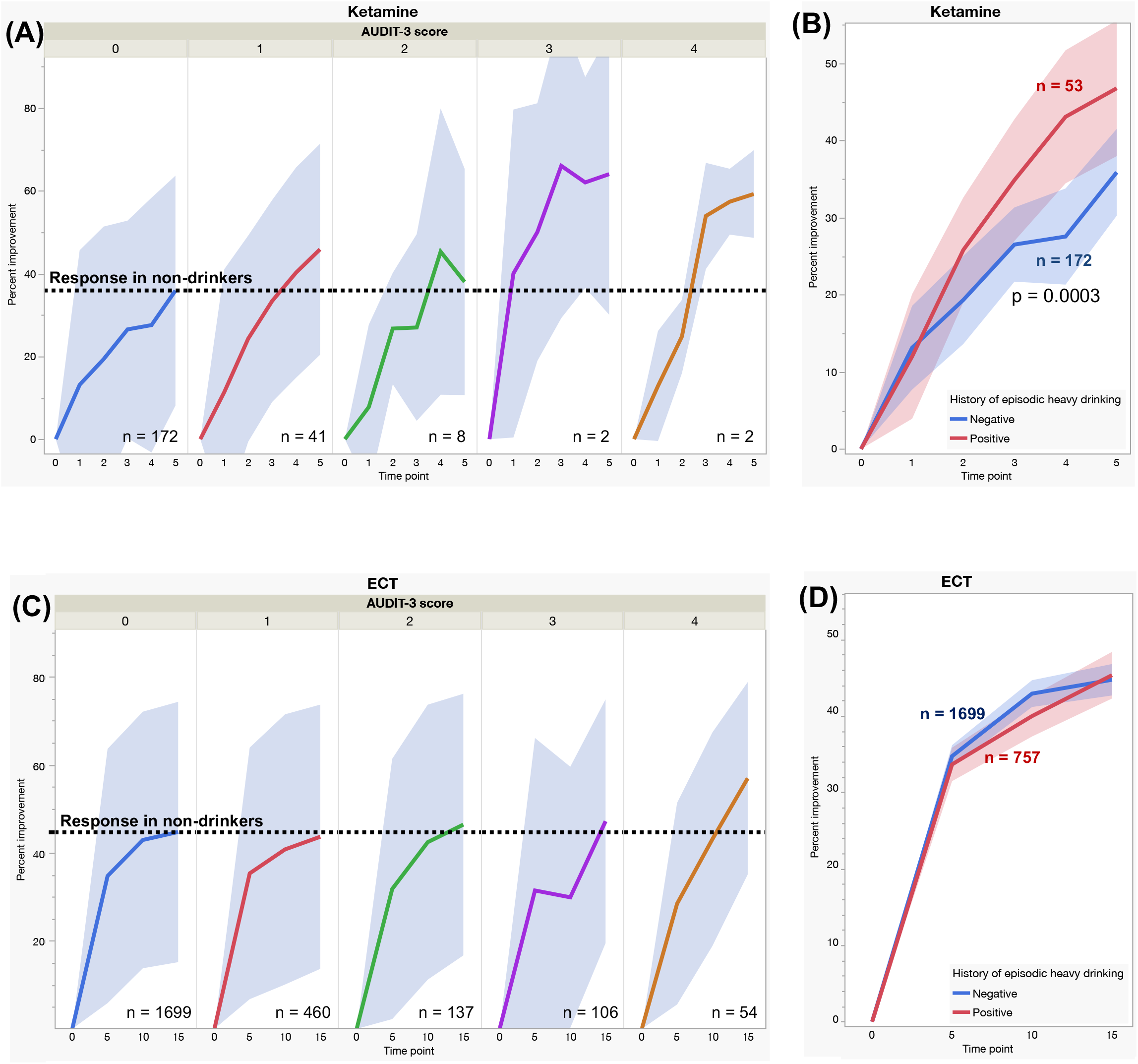
Prior episodic heavy alcohol use predicts antidepressant response to ketamine, but not ECT. **(a)** With ketamine, patients with a history of past-year episodes of heavy drinking (colored lines) showed greater percent improvement in depression from baseline relative to those without (black line) at treatments 3, 4, and 5. **(b)** Similar results were observed when binarizing history of alcohol use. **(c)** With ECT, past-year episodes of heavy drinking were generally not related to antidepressant outcome. Daily drinkers (AUDIT-3 = 4) appeared to respond better after treatment 15, but this did not survive multiple comparisons correction and was not seen when binarizing alcohol use **(d)**.

Antidepressant efficacy of ECT was not significantly predicted by AUDIT-3 score, as there was no significant group x time interaction on QIDS-SR scores after the first 5 treatments (F(4,2283)=0.91, p=0.46), 10 treatments (F(8,3939)=1.63, p=0.11), or 20 treatments (F(16,7154)=1.50, p=0.09) (Fig. 1c). Of note, there was an interaction after 15 treatments (F(12,5113)=1.93, p=0.027), although this is a post hoc result that is not corrected for multiple comparisons. This difference was also not detected when binarizing alcohol use history (Fig. 1d).

## Discussion

This retrospective cohort study revealed that patients who reported a higher frequency of past-year episodes of heavy drinking responded better to ketamine for treatment of trMDD than patients with a lower frequency of alcohol use. This effect was not seen in patients receiving ECT. This finding suggests that patients with AUD or those at higher risk for AUD may respond better to ketamine for depression. This result is consistent with prior studies documenting an association between those with a positive family history of AUD and antidepressant response to ketamine (Petrakis et al. 2004; Niciu et al. 2014), as these individuals are also at higher risk of developing AUD.

The association between AUD and effectiveness of ketamine may be related to the partly shared mechanisms of the two drugs. By contrast, ECT is not believed to work through opiate or NMDA receptors (NMDAR) (Fosse and Read 2013), which may explain why similar effects were not observed for ECT. Although alcohol’s acute effects are driven by GABAergic neurotransmission, the propensity towards AUD is believed to be mediated by its effect as an NMDAR antagonist(Krystal et al. 2003b), similar to the mechanism of ketamine (Orser et al. 1997; Krystal et al. 2003). There are several plausible mechanisms for this effect, although the direction of causality is unclear. For instance, if there is a subtype of depression that is more responsive to NMDAR antagonism, these patients may have been more inclined towards self-medicating with alcohol. Conversely, chronic alcohol use may cause depression that is more responsive to NMDAR antagonism via homeostatic upregulation of these and other glutamate receptors. It is also plausible that both phenomena are driven by shared genetics or other risk factors, as evidenced by the relevance of family history of alcoholism. Ketamine has also demonstrated some preliminary benefit as a treatment for AUD in the absence of MDD (Goldfine et al. 2023), which may further support the idea of a shared risk architecture.

These consults also may shed light into the ongoing quest to best allocate ketamine versus ECT in different contexts. A recent clinical trial-showed that ketamine was noninferior to ECT for nonpsychotic trMDD (Anand et al. 2023). While the ECT protocol used in this study may not have been optimal and there was a wide difference in dropout rate (Ekstrand et al. 2024), questions still remain regarding how to determine which treatment is optimal for a specific patient. Given that MDD and AUD are often comorbid (Sullivan et al. 2005; Castillo-Carniglia et al. 2019), a history of episodic drinking in the year before potential treatment may constitute one element considered in the evaluation of patients for treatment with ketamine. Although we did not directly compare ECT to ketamine in the present analysis due to many potential sources of selection bias, patients with no history of heavy drinking appeared to respond better to ECT, while those with some history of heavy drinking appeared to respond better to ketamine.

It is critical to note that ketamine is itself a substance of abuse with potential physical and mental health sequelae when used without close medical supervision (McIntyre et al. 2021). Ketamine has similarities to several addictive substances in structure, action, and effect including alcohol, opioids, phencyclidine, methylenedioxymethamphetamine, and others. Thus, it is plausible that ketamine treatment could potentiate or exacerbate alcohol use disorder in highly susceptible individuals. Further work is needed to refine whether specific symptoms or other features of alcohol use history affect the overall risk profile. Further studies are also needed to determine whether some patients may be at increased risk for adverse outcomes, such as ketamine use disorder. Given the increased availability of ketamine as a prescribed medication, care and attention should be paid to assessments, monitoring, patient education, and informed consent (McIntyre et al. 2021).

The retrospective nature of this study comes with limitations. A direct comparison between ECT and ketamine was not conducted as we would be unable to control for selection bias. For instance, the patients may differ in terms of treatment resistance, severity, and socioeconomic status amongst other variables. Similarly, there may be other differences between patients with and without a history of alcohol use, which may have affected other treatment-related parameters – for instance, patients with a history of AUD may tolerate higher doses of ketamine. Ketamine is also more novel and more expensive, which may lead to greater nonspecific effects such as expectation bias. It should also be noted that we have distinguished between Episodic Heavy Alcohol Use and Alcohol Use Disorder; full diagnosis of AUD is difficult to assess because patients with comorbid AUD have been either systematically excluded or underrepresented in many prospective studies of ketamine (Murrough et al. 2013; Daly et al. 2018). This trend also applies the present sample, which excluded patients who met formal criteria for an active substance use disorder in the ketamine group and did not systematically assess AUD diagnosis in the ECT group. As a result, only a small number of patients had very high AUDIT-3 scores; thus, trends within each AUDIT-3 subgroup were not analyzed due to inadequate power. Furthermore, ketamine may be less accessible to patients with AUD due to high out-of-pocket costs or required social and financial supports to attend treatments. Thus, patients with comorbid AUD are likely underrepresented in the current sample, suggesting that our reported effect is likely an underestimate. Future prospective studies, particularly those comparing ketamine to ECT, may further address this question by actively enrolling patients with and without comorbid AUD. Future studies may gain additional insights with sophisticated phenotyping, including remission status and duration, previous and concurrent recovery treatment activities, medication-assisted treatment, history of binge drinking versus daily drinking, length of time since last alcohol use, presence and nature of cravings, and other phenomenological aspects of AUD. More pronounced effects may also be detected with genotyping or assessment of synaptic plasticity with brain stimulation and/or NMDAR modulating drugs.

Although prospective studies are necessary, these retrospective results are strengthened by their consistency with prior clinical studies in patients with a family history of alcohol use disorder. Thus, these results suggest that personal history of alcohol use, in addition to family history, may better inform clinical treatment selection in patients who are considering ketamine treatment for MDD.

## Data Availability

Data from the present study may be shared upon reasonable request with an institutional data use agreement.

## Data Availability Statement

Due to the small number of patients treated in some of the groups described here, sharing patient-level data may present a risk to confidentiality. Thus, for this reason, we do not have IRB approval to share data.

## Acknowledgements

None.

## Ethical Approval

This study involved analysis of anonymized human data from an existing database and was thus determined by Mass General Brigham IRB to be exempt from formal review.

## Consent to participate

Not applicable. This study used fully anonymized data from an existing database; no identifiable information was accessible to the researchers, and no direct participant contact occurred.

## Consent to publish

Not applicable. This study used fully anonymized data from an existing database, and no identifiable individual data are included.

## Data Availability Statement

The data used in this study are not publicly available due to institutional privacy policies.

## Authors’ contributions

Conceived and designed the analysis: Robert C. Meisner MD, Shan H. Siddiqi MD, Aaishah Raquib MS Collected the data: Robert C. Meisner MD, Stephen Seiner MD

Curated the data: Robert C. Meisner MD, Aaishah R. Raquib MS, Vy Nguyen BS Performed the analysis: Shan H. Siddiqi MD

Wrote the paper: Aaishah R. Raquib MS, Vy Nguyen BS, Robert C. Meisner MD, Shan H. Siddiqi MD, with input from all authors

Provided content area expertise: Joshua C. Brown MD PhD, Stephen Seiner MD, Joseph J. Taylor MD PhD, Amanda E. Sedgewick DO, Robert C. Meisner MD

